## Supplementary Information Files for "Monitoring transmission intensity of trachoma with serology"

Tedijanto et al. 2023.

### Supplementary Information Materials File

**CONTENTS**

**Supplementary Table 1.** Overview of included studies.

**Supplementary Fig. 1.** Distribution of cluster sample sizes by study population.

**Supplementary Fig. 2.** Age distribution of children sampled by study population.

**Supplementary Fig. 3.** Relationship between study-level seroconversion rate estimates from models allowing for age-varying rates versus a constant rate.

**Supplementary Fig. 4.** Relationship between seroconversion rates and PCR prevalence

**Supplementary Fig. 5.** Identification of clusters with *C. trachomatis* infection using the seroconversion rate.

**Supplementary Fig. 6.** Identification of clusters with *C. trachomatis* infection using seroprevalence and the seroconversion rate among 1-5 year olds.

**Supplementary Fig. 7.** Robustness of seroprevalence and seroconversion rate to changes in seropositivity threshold.

**Supplementary Fig. 8.** AIC values for simple catalytic model, reversible catalytic models with a range of assumed seroreversion rates, and semiparametric spline (SS) model for each study-year.

### Supplementary Table 1. Overview of included studies.

| Country | Study population | Admin-istrative level | Number of clusters[1] | Number of children | Study type [2] | Year[3] | Trachoma program stage | Intervention context | Cita-tion |
|---|---|---|---|---|---|---|---|---|---|
| Ethiopia | Alefa | District | 21 | 704 | Survey | 2017 | From endemic to near elimination | Andabet, Dera, and Woreta town received MDA approximately 8 months prior to survey; Alefa had not received MDA for about 2.5 years | 1 |
| | Andabet | District | 21 | 569 | | | | | |
| | Dera | District | 22 | 628 | | | | | |
| | Woreta town | District | 11 | 261 | | | | | |
| Ethiopia | Wag Hemra (WUHA) | Area consisting of 3 contiguous districts | 40 | 2287 | RCT (NCT02754583) | 2019 | Endemic | 2-arm RCT compared immediate and delayed intensive water, sanitation and hygiene intervention; annual MDA conducted for 8 years prior, but suspended in both arms during RCT | 2 |
| Ethiopia | Wag Hemra (TAITU) | Area consisting of 3 contiguous districts | 37 | 1114 | RCT (NCT02754583) | 2018 | Endemic | 3-arm RCT compared MDA, triannual MDA to children 0–5-years-old with Ct infection, or delayed treatment; annual MDA conducted for 8 years prior to trial | 3 |
| Malawi | Chikwawa | District consisting of 3 EUs | 71 | 2630 | Survey | 2014 | Near elimination | MDA conducted annually from 2011 to 2013 | 4 |
| | Mchinji | District consisting of 3 EUs | 70 | 3271 | | | | | |
| Morocco | Agdaz | District | 28 | 1212 | Survey | 2019 | Post-validation | MDA stopped in 2005 | 5,6 |
| | Boumalne Dades | District | 29 | 1266 | | | | | |
| Niger | Dosso (MORDOR) | 2 districts within the Dosso Region | 29 | 939 | RCT (NCT02048007) | 2018 | Post-validation | RCT compared placebo to azithromycin distributed to children <5y in intervention arm to prevent mortality | 7,8 |
| Niger | Matameye (PRET) | District | 22 | 970 | RCT (NCT00792922) | 2013 | Endemic | 3-year 2x2 factorial RCT compared annual vs. 2x/year treatment and standard vs. enhanced coverage; serology collected only at final follow-up visit for enhanced coverage clusters | 9,10 |
| Tanzania | Kongwa | District | 8 | 1577 | RCT | 2013 | Near elimination | Area was highly endemic and received many rounds of MDA prior to study; RCT compared a single round of MDA at baseline to control; MDA was expanded in later study years | 11 |
| Tanzania | Kongwa | District | 50 | 2383 | Survey | 2018 | Post-validation | 2 years post-MDA | 12 |

*Abbreviations: RCT - randomized controlled trial; MDA - mass drug administration. EU – Evaluation Unit* Supplementary Table 1 footnotes: *1. Clusters with Pgp3 IgG measurements from at least 15 children available for this analysis*
*2. Randomized Controlled Trials (RCTs) also include clinicaltrials.gov protocol registration numbers (NCT…). The Kongwa 2013 trial was not formally registered with clinicaltrials.gov, but was overseen by IRBs at the US CDC and the Tanzanian Institute for Medical Research. 3. Study years included in this analysis. Some of the RCTs spanned multiple years. See Methods for details.*

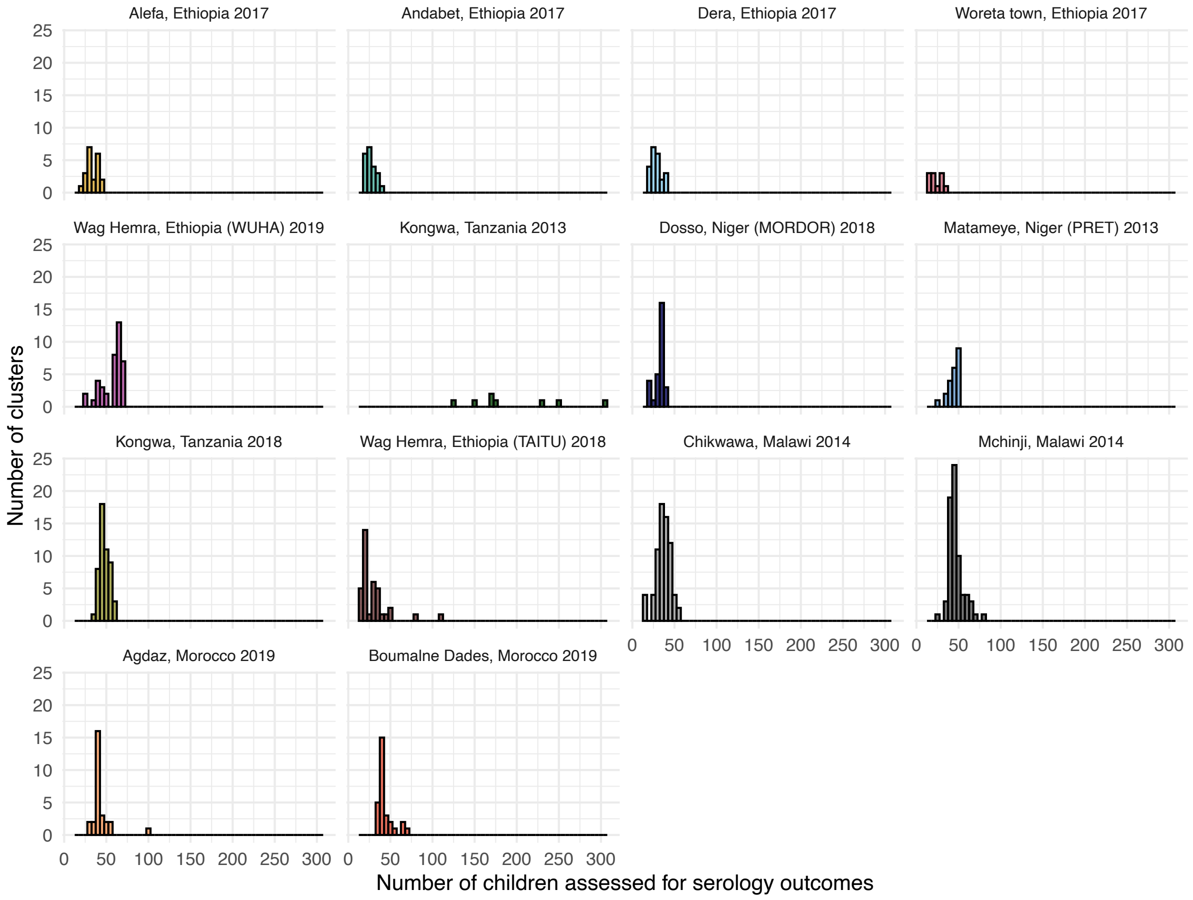


**Supplementary Fig. 1. Distribution of cluster sample sizes by study population.**
Created with notebook https://osf.io/qjst8 .

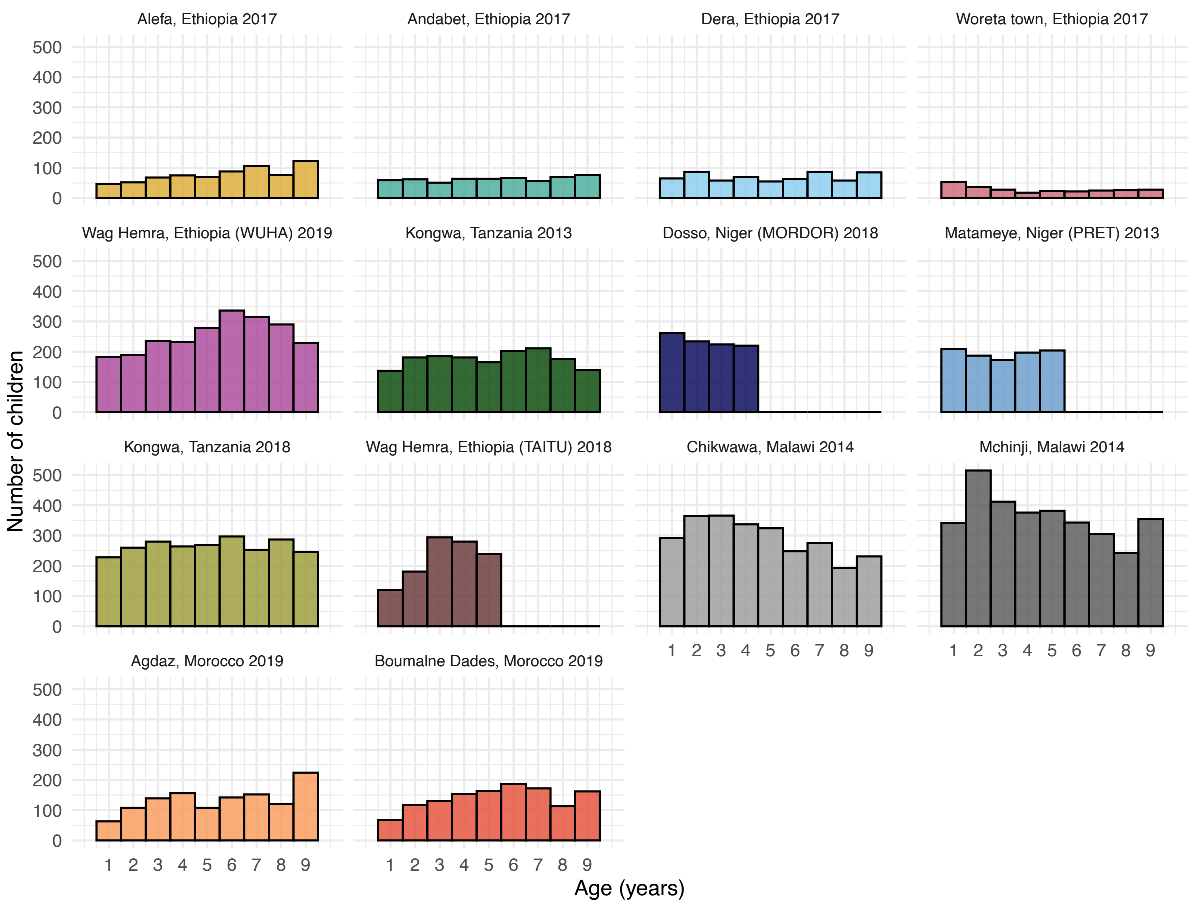


**Supplementary Fig. 2. Age distribution of children sampled by study population.** Created with notebook https://osf.io/qjst8 .

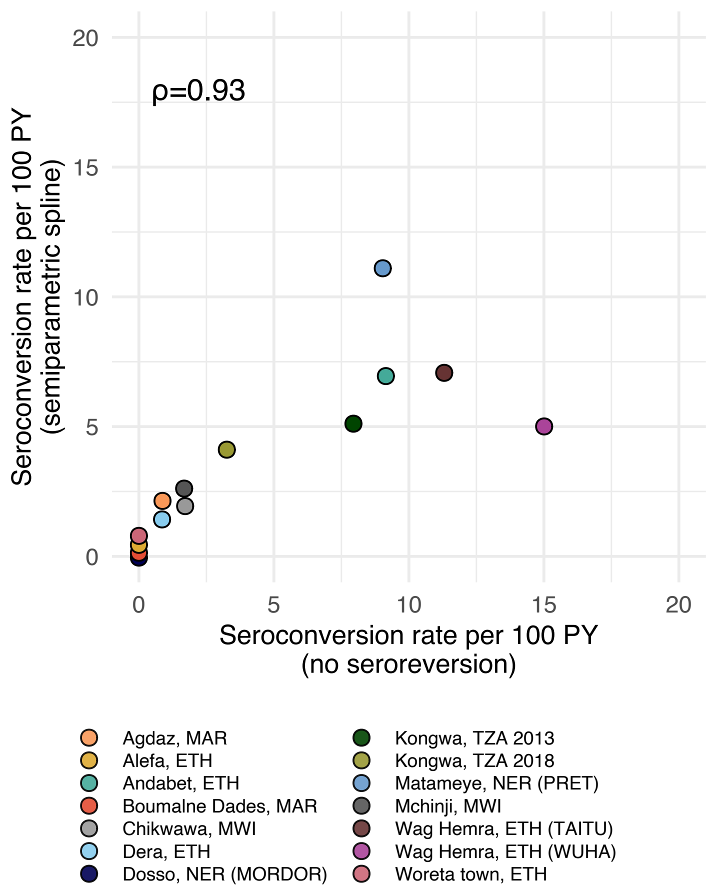


**Supplementary Fig. 3. Relationship between study-level seroconversion rate estimates from models allowing for age-varying rates versus a constant rate.** Estimates allowing for an age-varying rate used a semi-parametric spline to model age-seroprevalence and estimated an overall average rage over the age range (Methods). Constant rate model assumed no seroreversion. Spearman rank correlation is shown at top left. Due to sample size considerations, age-varying seroconversion rates using semiparametric spline were estimated at the study level only, as individual clusters included insufficient observations. Figure 1 of the main text includes sample sizes for each study. Abbreviations: ETH = Ethiopia; MAR = Moroccoo; MWI = Malawi; NER = Niger; TZA = Tanzania. Created with notebook https://osf.io/wcv86 .

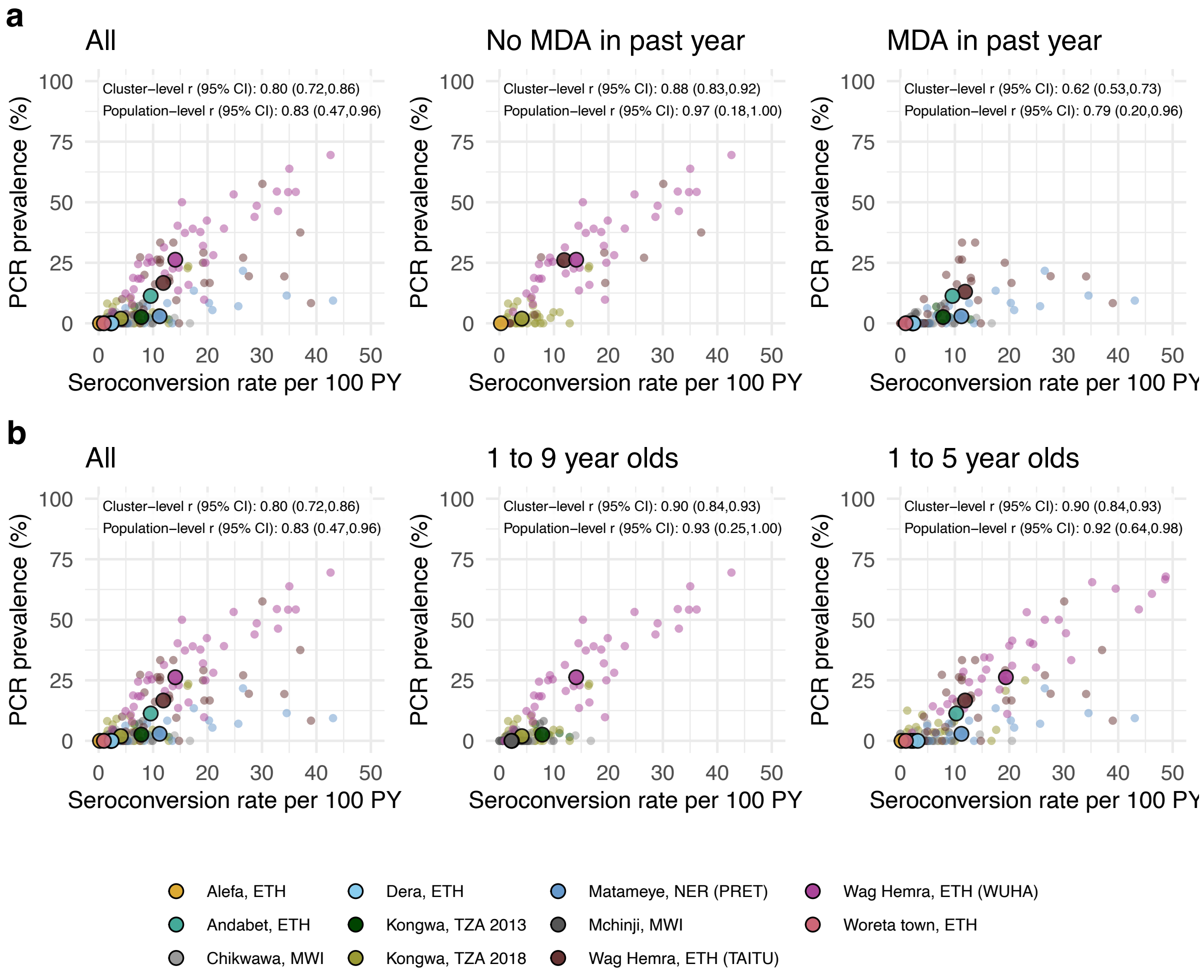


**Supplementary Fig. 4. Relationship between seroconversion rates and PCR prevalence.** **a** Comparison overall and stratified by whether the study population had received mass distribution of azithromycin (MDA) in the previous year. **b** Overall and stratified by child age, 1–9-year-olds compared to 1–5-year-olds**.** Medians across clusters for each study population are represented by larger points with black outline. Each plot includes the identity line (dotted) and Pearson correlations at the cluster- and population-levels. 95% confidence intervals (CIs) are based on 1000 bootstrapped samples, holding populations fixed and resampling clusters with replacement. Population-level values are included for Andabet, Dera, Woreta town, and Alefa, Ethiopia, but cluster-level PCR prevalence was not available for these populations. For some populations including 1–9-year-olds, measurements were already summarized at the population-level and prevalence could not be separately estimated among 1–5-year-olds. Abbreviations: ETH = Ethiopia; MWI = Malawi; NER = Niger; TZA = Tanzania. Created with notebook https://osf.io/rt825 .

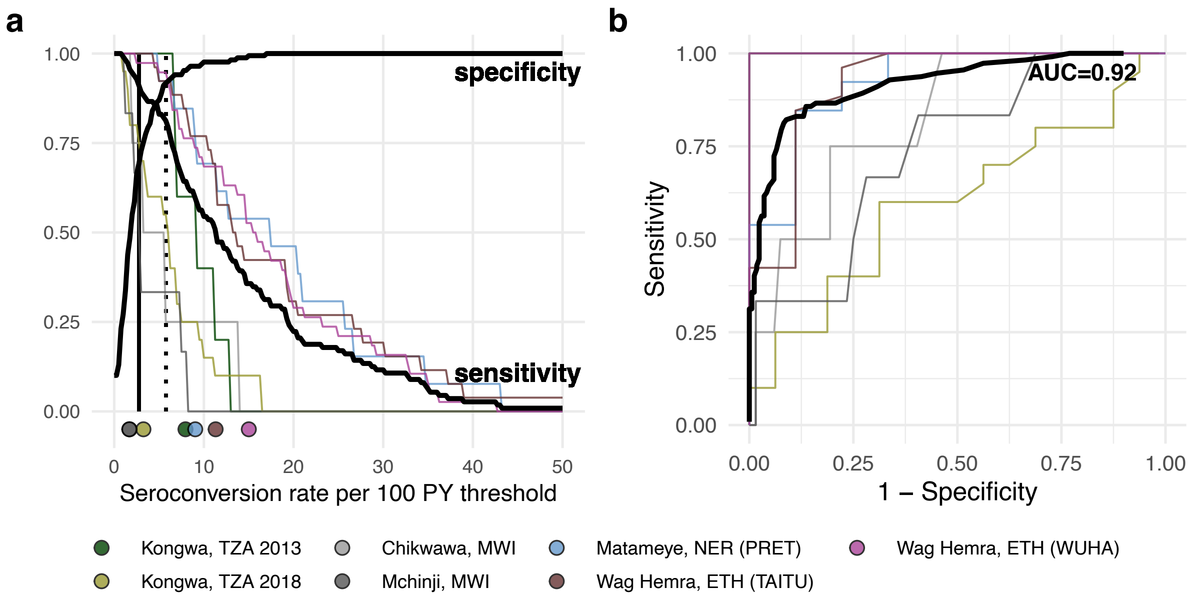


**Supplementary Fig. 5. Identification of clusters with *C. trachomatis* infection using the seroconversion rate**.

**a** Sensitivity and specificity for identification of clusters with any *C. trachomatis* infections based on different seroconversion rate thresholds among children 1-9 years old. Vertical lines delineate sensitivity at 90% (solid line, 2.75 seroconversions per 100 PY) and 80% (dotted line, 5.75 seroconversion per 100 PY). The thick lines represent overall sensitivity and specificity (n = 281 clusters; 112 clusters with PCR>0%), while thin lines show sensitivity for each study population. Points at bottom of plot mark seroconversion rate estimates for each study population. **b** Receiver operating characteristic (ROC) curves for all clusters (thick black line) and for each population (thin colored lines) with overall area-under-the-curve (AUC). Note that the ROC curve for Kongwa, TZA 2013 lies beneath the curve for Wag Hemra, ETH (WUHA). Abbreviations: ETH = Ethiopia; MWI = Malawi**;** NER = Niger; TZA = Tanzania**.** Supplementary Data 2 includes underlying sensitivity and specificity values at each cutoff. Source data are provided with this paper. Created with notebook https://osf.io/vgekh .

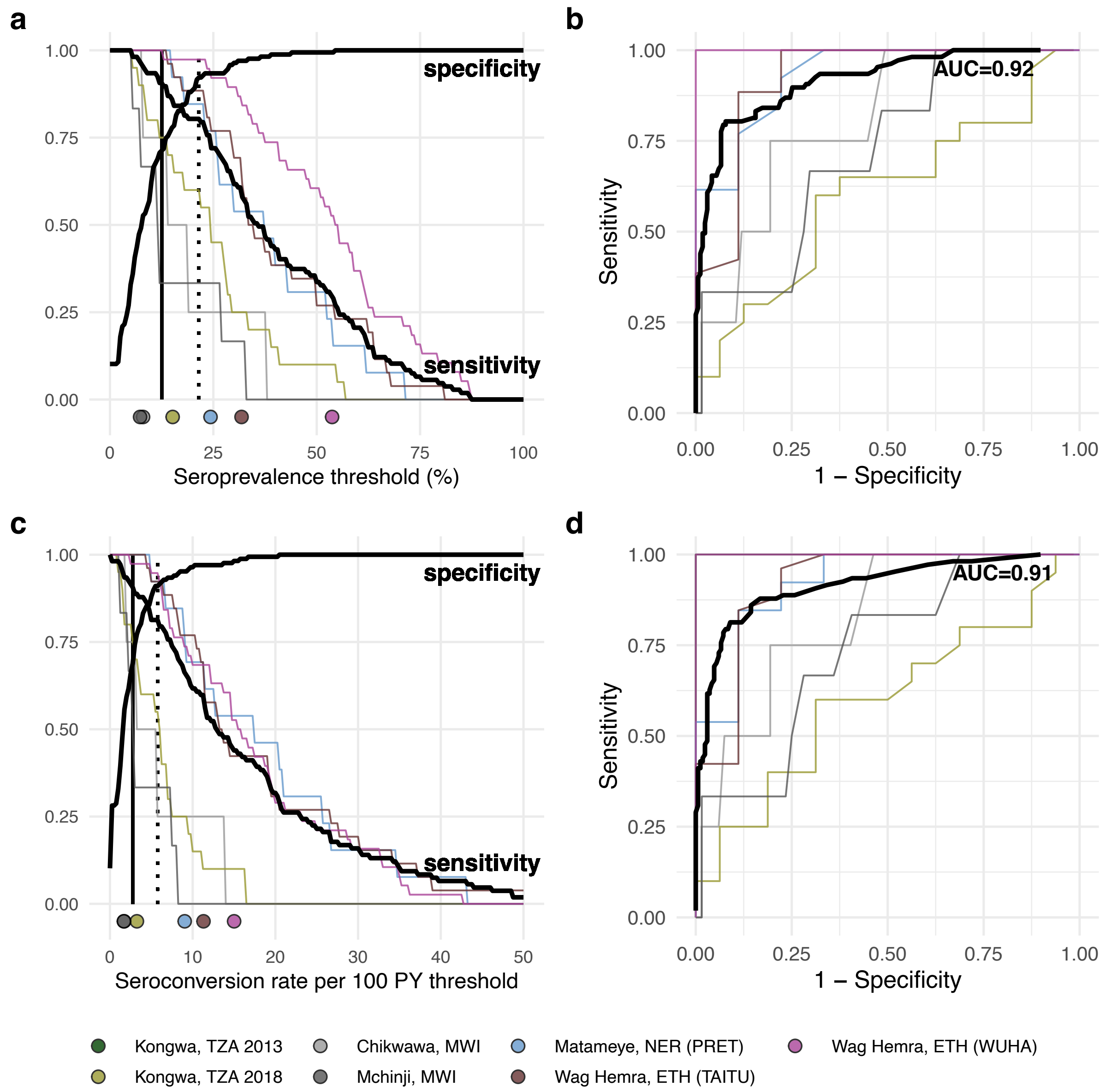


**Supplementary Fig. 6. Identification of clusters with *C. trachomatis* infection using seroprevalence and the seroconversion rate among children 1–5 years old**.
**a** Sensitivity and specificity for identification of clusters with any *C. trachomatis* infections based on different seroprevalence thresholds among children 1-5 years old. Vertical lines delineate sensitivity at 90% (solid line, 12.5% seroprevalence) and 80% (dotted line, 21.5% seroprevalence). The thick lines represent overall sensitivity and specificity (n = 274 clusters; 107 clusters with PCR>0%), while thin lines show sensitivity for each study population. Points at bottom of plot mark seroprevalence estimates for each study population. **b** Receiver operating characteristic (ROC) curves for all clusters (thick black line) and for each population (thin colored lines) with overall area-under-the-curve (AUC). **c** Estimates similar to panel a, but using the seroconversion rate. Vertical lines delineate sensitivity at 90% (solid line, 2.75 per 100 PY) and 80% (dotted line, 5.75 per 100 PY). **d** Estimates similar to panel b, but using the seroconversion rate. Note that the ROC curves for Kongwa, TZA 2013 in panels b and d lie beneath the curves for Wag Hemra, ETH (WUHA). Abbreviations: ETH = Ethiopia; MWI = Malawi**;** NER = Niger; TZA = Tanzania**.** Supplementary Data 3 and 4 includes underlying sensitivity and specificity values at each cutoff. Source data are provided with this paper. Created with notebook https://osf.io/vgekh .

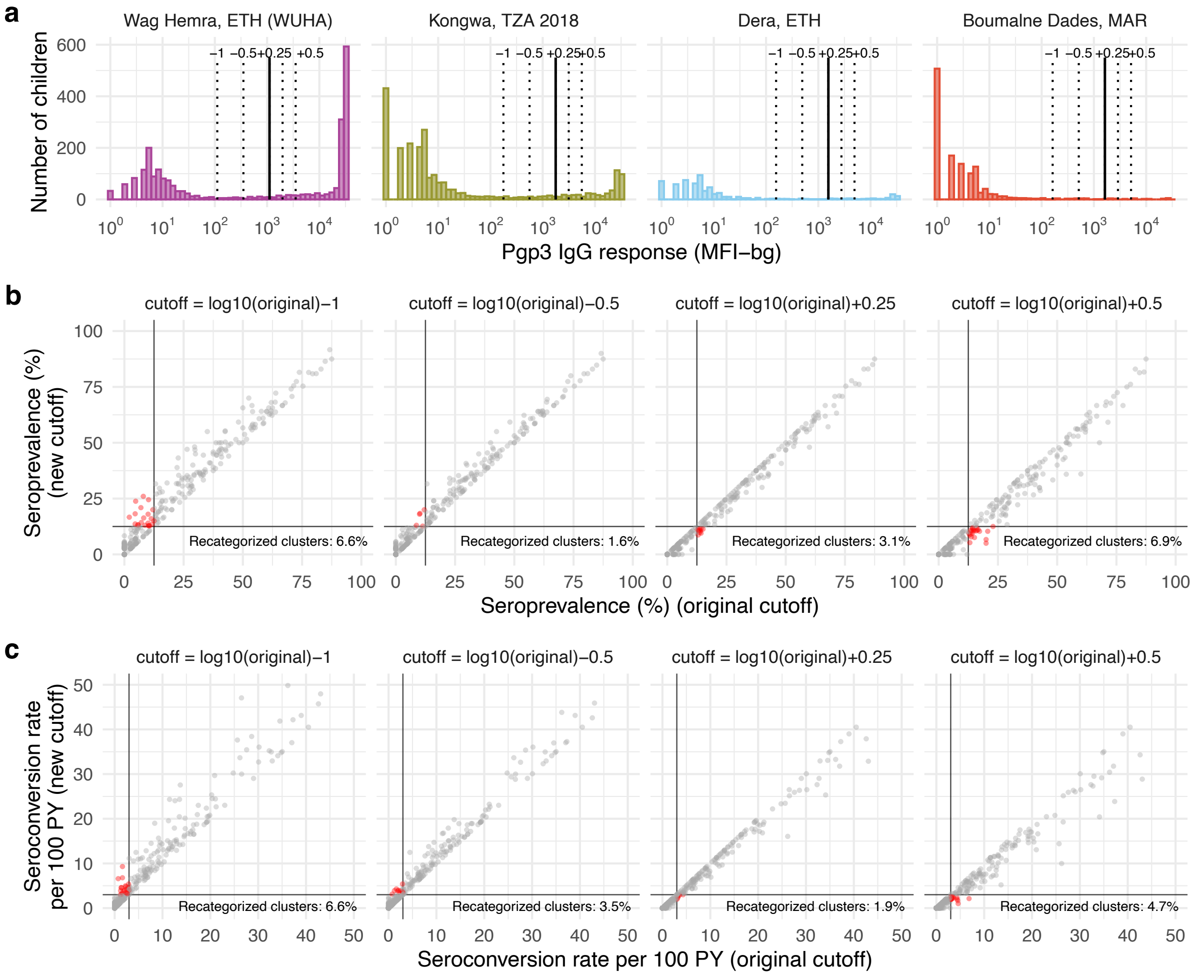


**Supplementary Fig. 7. Robustness of seroprevalence and seroconversion rate to changes in seropositivity threshold. a** Seroprevalence and seroconversion rate were recalculated using arbitrary seropositivity cutoffs at -1, -0.5, +0.25, and +0.5 in $\log_{10}$ units from the original cutoff, as shown in four populations representing a gradient of transmission settings. **b** At each new cutoff, we re-estimated seroprevalence and the proportion of clusters that were recategorized (red points) based on a threshold representing 90% sensitivity to identify clusters with any PCR infections (solid black lines, 13.5% seroprevalence, see Fig 6 in the main text). **c** At each new cutoff, we re-estimated the seroconversion rate assuming no seroreversion and the proportion of clusters that were recategorized (red points) based on a threshold representing 90% sensitivity to identify clusters with any PCR infections (solid black lines, 2.75 seroconversions per 100 PY, see Supplementary Fig 5). Abbreviations: ETH = Ethiopia; MAR = Morocco; TZA = Tanzania; PY = person-years. Source data are provided with this paper. Created with notebook https://osf.io/xs7nr .

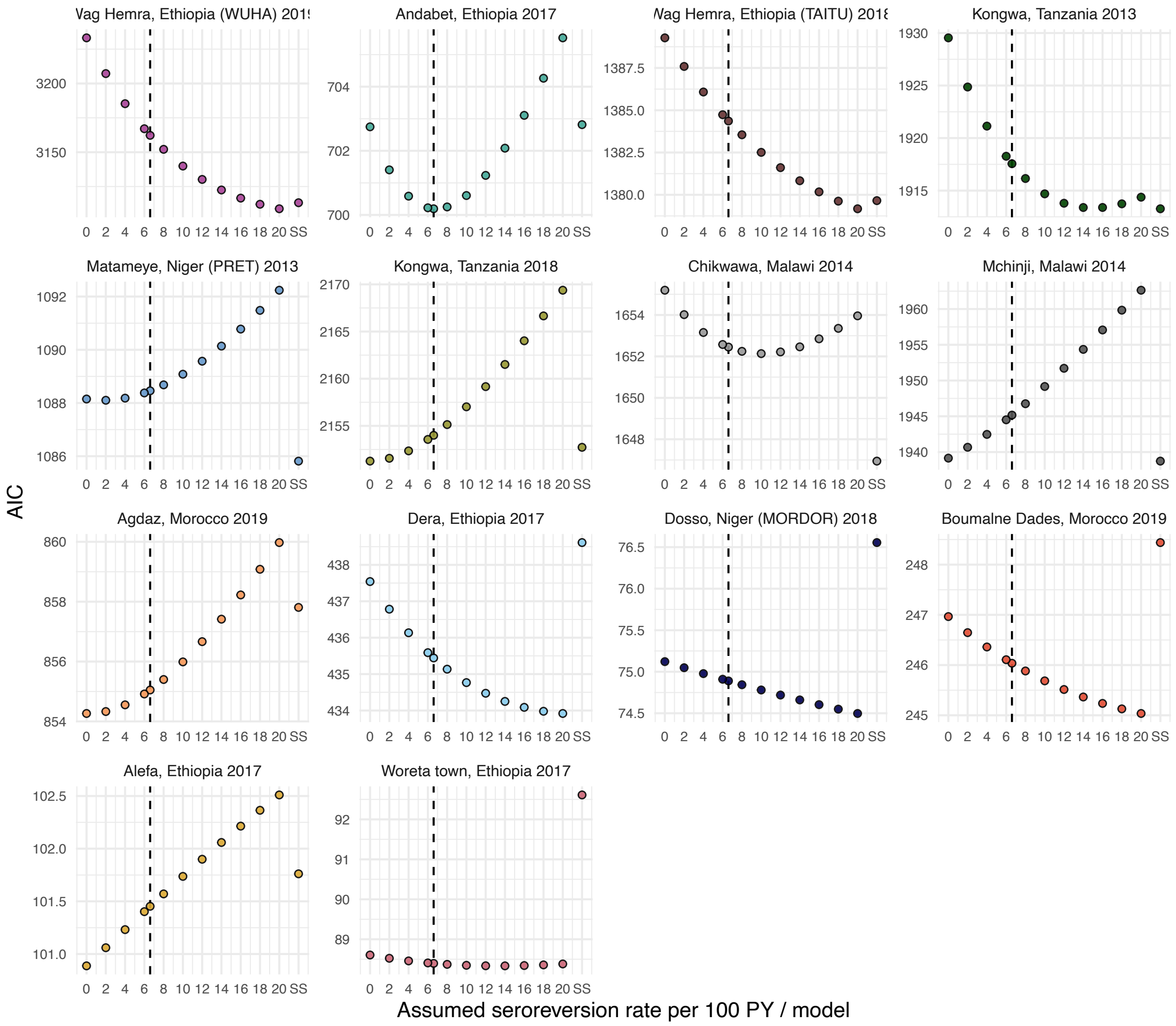


**Supplementary Fig. 8. Seroconversion model comparison based on AIC for catalytic models with and without seroreversion and semiparametric spline model.** In the main analysis, the catalytic model allowing for seroreversion assumed a seroreversion rate of 6.6 per 100 person-years and is marked with a vertical dashed line in each panel. The semiparametric spline model (SS) allows seroconversion rate to vary flexibly with age. Points may be missing in cases where models did not converge. Panels are arranged in order of descending population-level seroprevalence. Source data are provided with this paper. Created with notebook https://osf.io/wcv86 .
